# The gut microbiome and incident atherosclerotic cardiovascular events: Prospective analyses in three Swedish cohorts

**DOI:** 10.64898/2026.11.12.26367976

**Authors:** Kamalita Pertiwi, Ulf Hammar, Koen F. Dekkers, Eivind Coward, Oleg Kambur, Gabriel Baldanzi, Mario Delgado-Velandia, Sergi Sayols-Baixeras, Shuai Yuan, Peter Persson, David Gabrielsson, Mariam Miari, Camila Gazolla Volpiano, Guillaume Méric, Daniel Jönsson, Gunnar Engström, J. Gustav Smith, Karl Michaëlsson, Olle Melander, Johan Ärnlöv, Rob Knight, Robin Hofmann, Teemu Niiranen, Aki S. Havulinna, Veikko Salomaa, Kristian Hveem, Claes Ohlsson, Marju Orho-Melander, Susanna C. Larsson, Tove Fall

## Abstract

**Background:** Variation in the gut microbiome composition has been associated with atherosclerotic cardiovascular disease (ASCVD) in case-control designs. We aimed to investigate associations with incident ASCVD events applying a prospective design to reduce the impact of reverse causality.

**Methods:** We assessed associations between gut microbiome features measured by shotgun metagenomic sequencing and incident total ASCVD, ischemic heart disease (IHD) and ischemic stroke, including fatal and non-fatal events as defined by registry follow-up in 17,216 participants from three Swedish population-based cohorts (SCAPIS, SIMPLER, and MOS). Results from Cox proportional hazards regression models adjusted for demographic, and lifestyle factors, medication, and prevalent diabetes were combined across cohorts using a fixed-effect meta-analysis accounting for multiple testing. Associations were tested for replication in HUNT and FINRISK.

**Results:** During a median of 6-8 y follow-up, 769 incident ASCVD events occurred. Species diversity was inversely associated with ASCVD and IHD, with a hazard ratio for IHD of 0.86 per SD increase in species richness (95% CI: 0.79-0.95). Sixty-eight common species (i.e., present in ≥30% individuals) were associated with incident IHD after adjusting for sex and birth country, but only eight remained in fully adjusted models, including *Ruminococcus_B gnavus*. Among others, inverse associations were observed for SFEL01 sp004557245 and *Fimadaptatus* sp900553645 from *Christensenellales*, an order in which butyrate production has been reported. *Streptococcus mutans* and *Limosilactobacillus fermentum,* oral-associated species previously associated with coronary atherosclerosis, were associated with a higher risk of incident IHD, which remained after adjustment for dental health. Replication analysis in the HUNT study supported the finding for *S. mutans*. No associations were observed with ischemic stroke.

**Conclusions:** Gut species diversity and specific species abundance were associated with incident IHD, providing evidence supporting a protective role of two *Christensenellales* species and an adverse role of oral species and *R. gnavus* in the development of IHD.

## INTRODUCTION

The gut microbiome, a collection of microbes living in the gastrointestinal tract that plays important roles in human physiological processes, has been implicated in the pathogenesis of atherosclerotic cardiovascular disease (ASCVD).^1–5^ Proposed mechanisms for the role of the gut microbiome in ASCVD pathophysiology include the production of microbial-derived metabolites, such as short-chain fatty acids (SCFAs) and secondary bile acids, which are involved in lipid metabolism and immune signaling.^1,2^ Detection of bacterial DNA and biofilms of the oral pathogen *Streptococcus* in coronary plaques suggests an additional mechanism specific to oral bacteria. It has been postulated that oral bacteria and/or their components can translocate into the bloodstream as well as through the gastrointestinal tract, inducing local and systemic inflammatory responses, and modulating atherosclerosis.^3–5^

However, available human studies to date offer limited evidence of causality. Most published studies employed a case-control design and lacked control for confounding factors such as lifestyle and medications,^6,7^ known to affect the gut microbiome and modify ASCVD risk.^8–11^ Such designs cannot rule out the disease or other unaccounted factors causing the observed changes in the gut microbiome. Using data from 8,973 individuals without a history of CVD, we have previously reported gut enrichment of *Streptococcus* spp. associated with worse subclinical atherosclerosis in a cross-sectional analysis,^12^ providing support for the link between oral taxa and CVD risk. Longitudinal designs can help establish temporality; however, few studies have published follow-up data on ASCVD events, with largely null associations.^13–15^ These studies, which were mostly based on low-resolution 16S sequencing data that cannot reliably resolve bacterial species, included at most 254 events and thus were likely underpowered.

Microbial species associated with the development of ASCVD, therefore, remain incompletely characterized. We therefore set out to investigate the association between gut microbiome, measured by deep metagenomic sequencing, and the development of ASCVD in three large longitudinal Swedish cohorts including a total of 17,216 individuals free from ASCVD at baseline.

## METHODS

### Ethical considerations

The present study included data from three longitudinal Swedish cohorts (Fig. 1; discovery cohorts). The Swedish Ethical Review Authority has approved the conduct of the present study (DNR 2022-06137-01 with amendments 2023-04785-02, 2024-05416-02, and 2025-03082-02) and all included participants have provided informed consent. Approval was also obtained from the Regional Committee for Medical and Health Research Ethics Midt-Norge for the HUNT study (REK-656785) and the Coordinating Ethical Committee of the Helsinki and Uusimaa University Hospital District for the FINRISK cohort (558/E3/2001). FINRISK data were obtained with permission from THL Biobank and Findata. All studies were conducted according to principles laid out in the Declaration of Helsinki.

**Figure 1.**
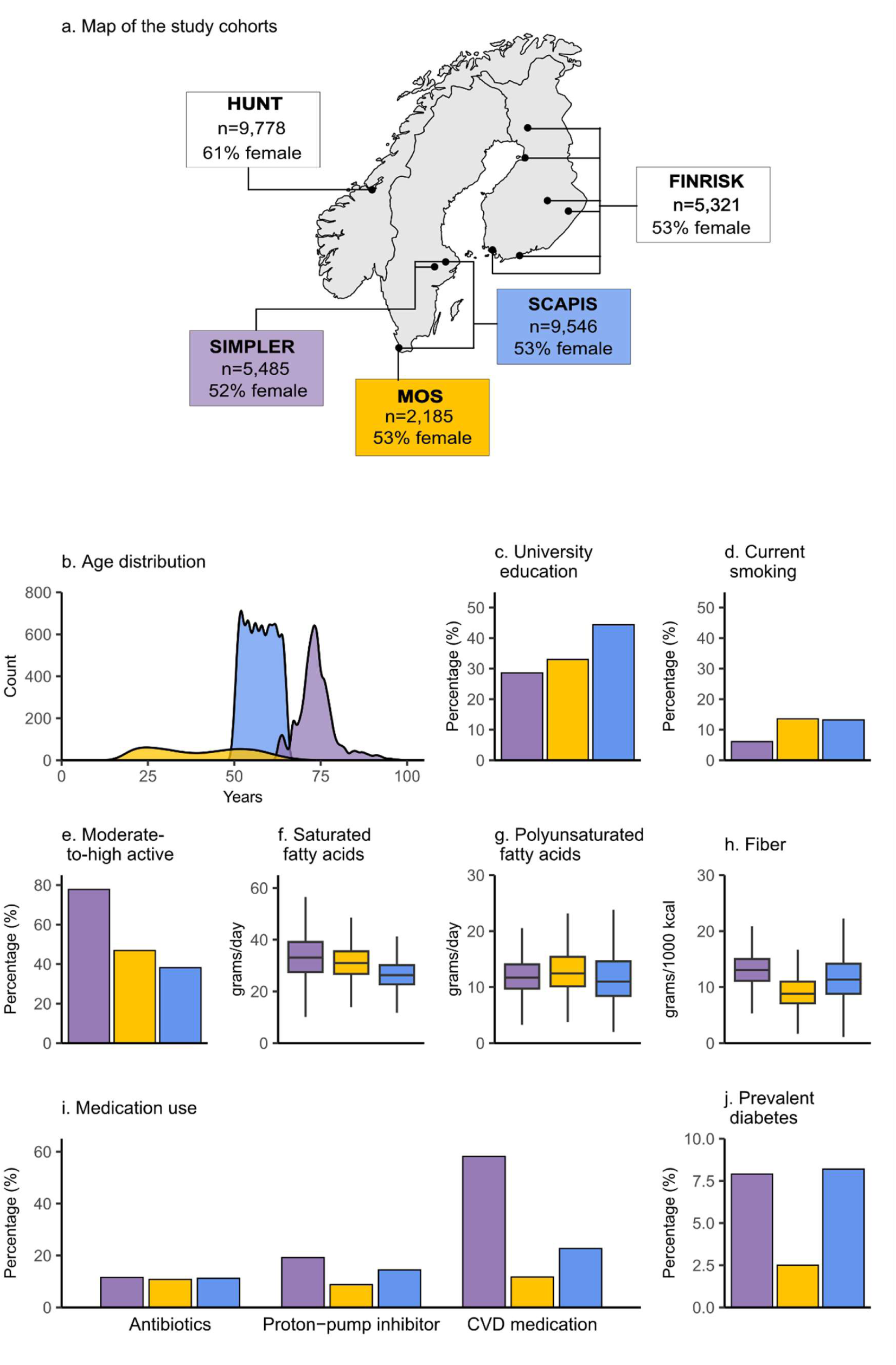
Study populations in the present analysis of gut microbiome and incident atherosclerotic cardiovascular disease. a) A map of countries of the cohorts included in the present analysis with dots on the map indicating study sites of each cohort, with sample sizes and proportion of females in the discovery (SIMPLER, SCAPIS, and MOS, indicated by purple, blue, and yellow-colored boxes, respectively) and the replication cohorts (HUNT and FINRISK, indicated by white boxes). b) Density plots show the age distribution of the discovery cohorts. Box plots show c) fiber, d) saturated and e) polyunsaturated fatty acid intakes in the discovery cohorts. Outlying data points are not shown. Bar plots show the proportion of individuals in the discovery cohorts who f) completed university education, g) were current smokers, h) engaged in moderate-to-high physical activity, i) had prevalent diabetes, and j) had recent antibiotic treatment, used proton-pump inhibitors, or cardiovascular medications at baseline. Values are also presented in Supplementary Table 1. CVD: cardiovascular disease; HUNT: Trøndelag Health Study; MOS: Malmö Offspring Study; SCAPIS: Swedish CArdioPulmonary BioImage Study; SIMPLER: Swedish Infrastructure for Medical Population-Based Life-Course and Environmental Research.

### Discovery cohorts

#### SCAPIS

The Swedish CArdioPulmonary BioImage Study (SCAPIS) aimed to improve understanding and prediction of cardiopulmonary diseases, and its design has been previously described.^16^ The participants were invited based on a random selection from the population register in six areas adjacent to six university hospitals across Sweden 2014-2018 (50% participation rate). A subset of 9,829 participants from Uppsala and Malmö sites collected fecal samples at home, stored at-20°C until the visit to the study site, then stored at-80°C until further processing.^17^

#### SIMPLER

The Swedish Infrastructure for Medical Population-Based Life-Course and Environmental Research (SIMPLER) cohorts comprise the Cohort of Swedish Men (COSM) and the Swedish Mammography Cohort (SMC).^18^ The two large ongoing population-based cohorts enrolled almost 110,000 individuals residing in Uppsala, Västmanland, and Örebro counties in 1997 (COSM) and 1987-1990 (SMC), respectively. During 2010-2011, a subsample of the participants from Uppsala and Västmanland was invited to a health examination and was asked to collect fecal samples at home and store the samples in home freezers (-20°C) and bring them to the study visit, except for a few participants in Uppsala who received home visits (*n*=196). The fecal samples were then stored at-80°C until further processing.

#### MOS

The Malmö Offspring Study (MOS) was established to study the influence of family-related factors on the development of chronic diseases.^19^ Adult children and grandchildren (≥18 y) of the participants of the cardiovascular arm of the Malmö Diet and Cancer study were invited to participate in the MOS. Consequently, MOS includes related participants. Between 2013 and 2017, 2,644 participants were recruited. The participants were asked to collect fecal samples after the first visit and store them at home (-20°C) until the second visit to the study site, after which the samples were stored at-80°C. The present study included participants who were not part of the SCAPIS cohort.

### Population for analysis – discovery cohorts

For the present analyses, only participants with fecal metagenomic data passing quality control and who consented to registry linkage were eligible (SCAPIS: 9,791; SIMPLER: 6,150; MOS: 2,215). Because we aimed to investigate new events of ASCVD, data from participants with a history of ischemic heart disease (IHD) or ischemic stroke at fecal sampling were excluded. This resulted in a total of 17,216 individuals free from ASCVD for analysis (SCAPIS: 9,546; SIMPLER: 5,485; MOS; 2,185 (Supplementary Fig. 1)).

### Metagenomic DNA extraction and shotgun metagenomic sequencing

DNA extraction from fecal samples and subsequent metagenomic sequencing in SCAPIS, SIMPLER, and MOS cohorts have been previously described in detail.^12,17^ For SCAPIS and MOS samples, DNA extraction, quality control, and sequencing were performed by Cmbio (Copenhagen, Denmark). In short, at least one negative control and positive control (ZymoBIOMICS™ Microbial Community Standard (D6300; Zymo Research)) were included in each batch starting from DNA extraction. The DNA was extracted from the fecal samples by using the NucleoSpin 96 Soil extraction kit (Macherey–Nagel, Germany), quantified with fluorometric techniques, and fragmented. DNA library construction was performed using the NEBNext Ultra Library Prep Kit from Illumina. Sequencing was performed using the Illumina Novaseq 6000 Instrument with 2×150-base-pair (bp) paired-end reads, resulting in an average of 26.0 and 25.3 million read pairs/sample for SCAPIS and MOS, respectively. Phred quality score >20 was observed for 97.8% of the sequenced bases in both SCAPIS and MOS.

Metagenomic DNA extraction and sequencing for SIMPLER was performed by the Centre for Translational Microbiome Research at the Karolinska Institutet in Stockholm, Sweden. Stored fecal samples from SIMPLER participants were thawed and a pea-sized aliquot was prepared to which DNA/RNA Shield (Zymo Research) was added, and the samples were then refrozen in the biobank before delivery, with each batch containing one negative and one positive control. DNA extraction from samples was performed using MagPure Stool kit (Magen Biotechnology), and the DNA was subsequently fragmented and used to generate the library using the MGIEasy FS DNA Library Prep Set kit. The sequencing was performed using 2×150bp paired-end reads on the DNBSEQ G400 or T7 sequencing instrument (MGI), resulting in an average yield of 50.8 million read pairs/sample.

### Taxonomic profiling

Identified host read pairs based on the mapping to the human reference genome were removed using Bowtie2 v.2.4.2 (SCAPIS and MOS) or Kraken 2 (SIMPLER). Subsequent bioinformatic processing, including calculation of relative abundances and annotation to taxonomic ranks, was performed by Cmbio (Copenhagen, Denmark) using the CHAMP profiler,^20^ based on the Human Microbiome Reference HMR05 catalog^20^ and the Genome Taxonomy Database (GTDB) release 214 (date: 28 April 2023).^21,22^ In non-rarefied data, 4,741 species were detected in SCAPIS, 3,927 in MOS and 4,676 in SIMPLER. Species richness, Shannon index, and inverse Simpson index were calculated as measures of alpha diversity on rarefied data, with the latter two indices calculated using the vegan R package (R v4.3.1).

The main exposures in the present study were alpha diversity, and relative abundances of metagenomic species present in at least 30% of individuals in each discovery cohort, resulting in 427 species, hereafter referred to as “common gut species.” Additionally, we examined an additional 114 rare species (<30% prevalence) hereafter referred to as “rare candidate species,” that were associated with coronary artery calcium score (CACS) in SCAPIS (Supplementary Methods) using the same model specification as in the previous SCAPIS publication.^12^

### Ascertainment of ASCVD events

The three primary outcomes were 1) incident total ASCVD, which included IHD and ischemic stroke events, and separately, 2) incident IHD, or 3) incident ischemic stroke. Information about ASCVD events was obtained through linkage to the Swedish National Patient Register and the Cause of Death Register. Individuals with prevalent ASCVD at baseline were identified and excluded from all association analyses. IHD included non-fatal and fatal IHD. Non-fatal IHD included ICD-10 codes I21-I22 or ICD-9 410 as main diagnoses, or coronary procedures such as percutaneous coronary intervention and coronary artery bypass graft surgery (Nordic surgery codes FNG02, FNG05, FNC, FND, and FNE). Fatal IHD included deaths coded as I20-I25 as the underlying cause of death. Ischemic stroke was defined as events with ICD-10 I63 or ICD-9 433-434 as main diagnosis or underlying cause of death. For analysis with total ASCVD as the outcome, only the first occurrence of either IHD or ischemic stroke was considered. Follow-up time was calculated for each individual from the date of fecal sampling (for SIMPLER) or screening date (for SCAPIS and MOS) until the date of incident event, death, emigration date, or the end of follow-up, whichever came first. The end of follow-up was 31 December 2022 for SIMPLER, 31 December 2023 for MOS, and 30 September 2024 for SCAPIS.

### Covariates

Covariates are harmonized across discovery cohorts when possible; expanded information is provided in the Supplementary Methods. Information on age and sex was derived from the Swedish Population Register. Education and physical activity level were self-reported by using questionnaires. Education level was categorized into ≤9 years, 10-12 years, and >12 years. Physical activity was limited to leisure time, with cohort-specific categories. Birth country, based on questionnaire responses or linkage to the Swedish Tax Register, was categorized in SIMPLER and SCAPIS into Sweden, Europe other than Sweden, and others, and in MOS as Sweden or others. Smoking status was categorized into current, former, or never smokers. Data on usual dietary intakes were collected at the same time as fecal sampling, using validated food frequency questionnaires in SCAPIS^23^ and SIMPLER,^24^ or a 4-d web-based food record in MOS,^25^ and nutrient intakes were obtained by linkage to the Swedish food composition database. Information on medication use across all cohorts was obtained primarily through linkage to the National Prescribed Drug Register and was additionally confirmed by questionnaire or detected drug metabolites in blood when available.

### Cardiovascular risk factors

During visits to study centers, participants underwent anthropometric measurements including height (in cm) and weight (in kg), from which body mass index (BMI, kg/m^2^) was calculated, and waist circumference (in cm). Systolic and diastolic blood pressure were measured twice, and the average of the two measurements was used. Venous blood samples were collected (non-fasted samples <1% in SCAPIS). Fresh plasma samples were used to measure glucose (mmol/L) with the enzymatic method with hexokinase, lipids (total cholesterol and HDL cholesterol (HDL-C), mmol/L), high-sensitivity C-reactive protein (hsCRP, mg/L) using the immunoturbidimetric method, and creatinine (µmol/L) with the enzymatic colorimetric method. LDL cholesterol (LDL-C, mmol/L) was directly measured or calculated using Friedewald’s formula. EDTA-plasma samples were used to measure apolipoprotein B (ApoB), lipoprotein(a) (Lp(a)), cystatin C (mg/L), and liver enzymes aspartate aminotransferase (ASAT, µkat/L), alanine aminotransferase (ALAT, µkat/L), gamma-glutamyltransferase (GGT, µkat/L). Estimated glomerular filtration rate (eGFR) was calculated using the CKD-EPI 2021 formula using both creatinine and cystatin C.^26^

### Plasma metabolites

The procedure has been described in detail previously in Dekkers et al.^27^ In short, the measurement and identification of plasma metabolomes in SCAPIS was performed by Metabolon (Durham, NC, USA). The EDTA plasma samples were analyzed in three different batches: Malmö (*n*=3,720), Uppsala-1 (*n*=2,825), Uppsala-2, (*n*=1,855). Untargeted ultrahigh-performance liquid chromatography-tandem mass spectrometry (UPLC-MS/MS) across four different platforms was used to measure the metabolites, which were subsequently identified by comparison with a reference standard library. When the measured value was below the detection threshold, the next higher value was used to impute the measured value.

### Dental health measures

Data on dental procedures, diagnoses, and the number of teeth were obtained for 96% SCAPIS participants through linkage to data from the Swedish National Dental Health Register (during 2010-2020) and were processed to derive measures of periodontitis status, caries status, and the number of missing teeth. The derivation and validation of these measures have been described in detail previously.^28,29^ Severe periodontitis status (present vs. absent) was defined by using records indicating a history of tooth extraction or surgical treatment (procedural codes starting with “4”) due to periodontitis (condition “periodontitis”) and two extensive sessions of nonsurgical periodontal treatment (procedural codes 342/343, along with the condition “periodontitis”). Caries status was defined as no entries (no caries) or any entries (caries) due to primary or secondary caries (any procedure paired with condition code 4001, 4002, 4011, 4012, or 4022). For descriptive purposes, the number of missing teeth was also categorized as ≥5 or <5.

### Replication cohorts

#### HUNT

The Trøndelag Health Study (the HUNT study) is an ongoing population-based cohort comprising adults ≥20 y residing in Trøndelag county, Norway, and has recruited >200,000 participants since its start in 1984. Between 2017 and 2019, the participants were invited to the HUNT4 survey, and 56,042 individuals were included.^30^ Of these participants, 13,268 participants provided fecal samples for gut microbiome measurement. For the replication analysis, microbiome data were available from 12,887 participants. After excluding 2,736 participants with missing covariate data and 373 prevalent IHD cases, data from 9,778 participants were analyzed. Median follow-up was 5.4 y. Methods for gut microbiome measurement, outcome and covariate definition are further described in the Supplementary Methods.

#### FINRISK

The FINRISK cohort is a national cohort initiated in 1972 whose data collection was conducted every 5 years by the Finnish Institute for Health and Welfare up to 2012.^31^ The data used in this study were from the FINRISK 2002 cohort, which included 8,738 participants aged 25-74. At the 2002 baseline, the participants completed questionnaires on sociodemographic and lifestyle factors, underwent health examinations, and donated fecal samples. For the replication analysis, participants without fecal samples, with missing covariates or with prevalent IHD were excluded (total *n* excluded=1,722), resulting in data from 5,321 individuals, with a median follow-up of 17.8 y. Methods for gut microbiome measurement and definition for the outcome and covariates are further described in the Supplementary Methods.

## Statistical analysis

### Association of gut microbial features and incident ASCVD

A base-2 logarithmic function was used to transform non-rarefied species relative abundance data after replacing zero values with half of the species-specific minimum value. Cox proportional hazards models with age as the underlying timescale were used to estimate hazard ratios and 95% confidence intervals (HR [95% CI]) for associations between gut microbial features and incident total ASCVD, IHD, and ischemic stroke, implemented using the survival R package (v.3.5-5).

Two analytical models were used. The minimally adjusted model included sex, birth country, and a technical covariate (metagenomic extraction plate in SCAPIS and MOS, aliquoting plate in SIMPLER). The fully adjusted model additionally included education level, smoking status, leisure time physical activity, energy-adjusted intakes of carbohydrate, polyunsaturated fatty acid, saturated fatty acid, and fiber, total energy intake, alcohol intake (not energy-adjusted), cardiovascular medication use, proton-pump inhibitor use, and prevalent diabetes. To account for the relatedness among MOS participants, we added a frailty term to the Cox models for MOS data. All covariates were specified *a priori*, and a directed acyclic graph was used to inform the adjustment sets (Supplementary Fig. 2). To handle missing data in covariates, multiple imputation with ten imputed datasets was performed in each discovery cohort, and estimates from the imputed datasets were pooled using Rubin’s rules; all implemented using the mice R package (v3.16.0). Details for missing data handling were further expanded in Supplementary Methods. All analyses were performed using RStudio v2023.12.1.402 (R v4.3.1), unless specified otherwise.

### Meta-analysis of discovery cohorts

Cohort-specific estimates were meta-analyzed using a fixed-effects model with inverse-variance weighting to estimate outcome-specific hazard ratios and 95% CIs for each microbiome feature (alpha diversity and metagenomic species). Microbiome features were considered to be associated with the outcome in the meta-analysis when passing multiple testing correction using the Benjamini-Hochberg false discovery rate set at 5% (*q*<0.05). For the outcome total ASCVD and IHD, cohort estimates from 80 rare candidate species with prevalence >1% in SIMPLER and SCAPIS, and >10% in MOS (for total ASCVD only) were meta-analyzed due to unstable estimates for species below these prevalence cut-offs. For the outcome ischemic stroke which had much fewer events, we used the cut-off prevalence of 1.5%, and estimates from 75 species were meta-analyzed. Meta-analyses were performed using the metafor R package (v4.8.0) in RStudio v2025.05.0+496 with R v4.5.2.

### Sensitivity analysis

Sensitivity analyses were performed in each cohort, and results from each cohort were subsequently meta-analyzed. We 1) further adjusted the results from the primary analyses (i.e., the fully adjusted model) for BMI, 2) excluded individuals receiving antibiotic treatment six months prior to fecal sampling, 3) excluded those with prevalent inflammatory bowel diseases, and 4) excluded the first two years of follow-up by starting the follow-up time two years after the fecal sampling to assess potential reverse causality. For analysis of microbial species, we also 5) further adjusted the results for richness, 6) performed censoring after three years of follow-up, and 7) applied different types of transformations to relative abundance data (details in Supplementary Methods). In SIMPLER, collection of fecal samples occurred over a longer time span relative to SCAPIS or MOS; we therefore also performed an additional sensitivity analysis to examine this effect by 8) including age-at-entry as a stratifying variable when fitting Cox regression on SIMPLER data.

### Correlation of microbiome features with risk factors

We performed partial Spearman correlations for microbial features identified in the primary analyses with age, sex, birth country, adiposity (BMI, waist circumference), and clinical ASCVD risk factors when available in the cohort, including plasma glucose, lipids, hsCRP, and measures of kidney and liver function. For age, sex, and birth country, correlations were adjusted for technical variation (plate). For all other risk factors, Spearman’s partial correlations were adjusted for age, sex, and technical variation. Multiple testing adjustment was performed using a 5% false discovery rate per microbiome feature identified in the primary analyses.

### Correlation of microbiome features with plasma metabolites

We performed partial Spearman correlations between microbial features identified in the primary analyses and 1,326 plasma metabolites in the SCAPIS cohort (max *n*=8,400), adjusting for metabolomics delivery batch, metagenomic extraction plate, age, sex, and birth country. We analyzed all metabolites detected in at least 100 samples. For metabolites that were detected in <2% of the individuals or classified as drug metabolites (*n*=105), these were analyzed as binary variables (detected/not detected), with the exception of 2,6-dihydroxybenzoic acid and salicylate (detected in >99% of the individuals). Multiple testing adjustment was performed using the 5% false discovery rate per microbiome feature. To investigate potential shared metabolic pathways among microbial features identified in the primary analyses, we further selected metabolites with moderate correlations (absolute correlation≥0.3) with richness or the IHD-associated species. Metabolites with the 20 strongest correlations were selected instead when the strongest correlation with a species was <0.3.

### Post-hoc analysis for associations with dental health

We additionally adjusted the Cox models from the primary analysis for measures of dental health, one at a time.

### Replication in HUNT and FINRISK

Microbiome features passing the multiple testing correction threshold in the fully adjusted model (*q*<0.05) were analyzed in the replication cohorts (HUNT and FINRISK) using the same model specification as in the discovery cohorts. Details on replication cohorts are provided in the Supplementary Methods. Findings were considered replicated when associations in the discovery and replication cohorts were in the same direction and were statistically significant using two-sided tests (*P*<0.05).

## RESULTS

We analyzed fecal metagenomic data from 17,216 individuals without a history of ASCVD across three large longitudinal Swedish cohorts (Figure 1, Supplementary Table 1). The SIMPLER cohort comprised older individuals than those in SCAPIS and MOS; it also had fewer current smokers, and a higher proportion of individuals using cardiovascular medication and proton-pump inhibitors at the time of fecal sampling. During up to 10 y follow-up, we identified 769 ASCVD events, largely due to IHD (514 events; Supplementary Table 2). For IHD and ischemic stroke, estimates were available only from the two larger cohorts, as only 22 IHD and 9 ischemic stroke events occurred in MOS.

Individuals who developed IHD events during the follow-up were more often males, and at baseline, less likely to be never smokers or be physically active, had higher saturated fatty acid and lower fiber intakes, as compared to those who remained free from IHD. Those who later experienced IHD events were also more likely to have diabetes and had a worse cardiovascular risk profile at baseline, despite also being more likely to be using cardiovascular medications (Supplementary Table 3). Individuals with higher microbial alpha diversity, measured by species richness at baseline, were more likely to be female, highly educated, more physically active, never smokers, and to have a healthier diet (i.e., higher fiber and lower saturated fatty acid intakes), but less likely to have diabetes or used antibiotics recently (Supplementary Table 4).

### Associations of alpha diversity with ASCVD

Higher microbial diversity was associated with a reduced hazard of incident ASCVD events, with estimates attenuated slightly after accounting for lifestyle, diet, medication use and presence of diabetes in the fully adjusted model (e.g., for Shannon, HR_full_=0.92 [0.85-0.99]) (Fig. 2, Supplementary Table 5) and with incident IHD (e.g. a 13% reduction of IHD hazard per 100-species increase in richness (HR_full_=0.87 [0.79-0.95]). Results were similar when excluding those with recent antibiotic use, or prevalent inflammatory bowel disease, or when excluding the first two years of follow-up (Supplementary Fig. 3), or when adjusting for BMI (Supplementary Fig. 3, Supplementary Table 13). No associations were observed for ischemic stroke (Fig. 2).

**Figure 2.**
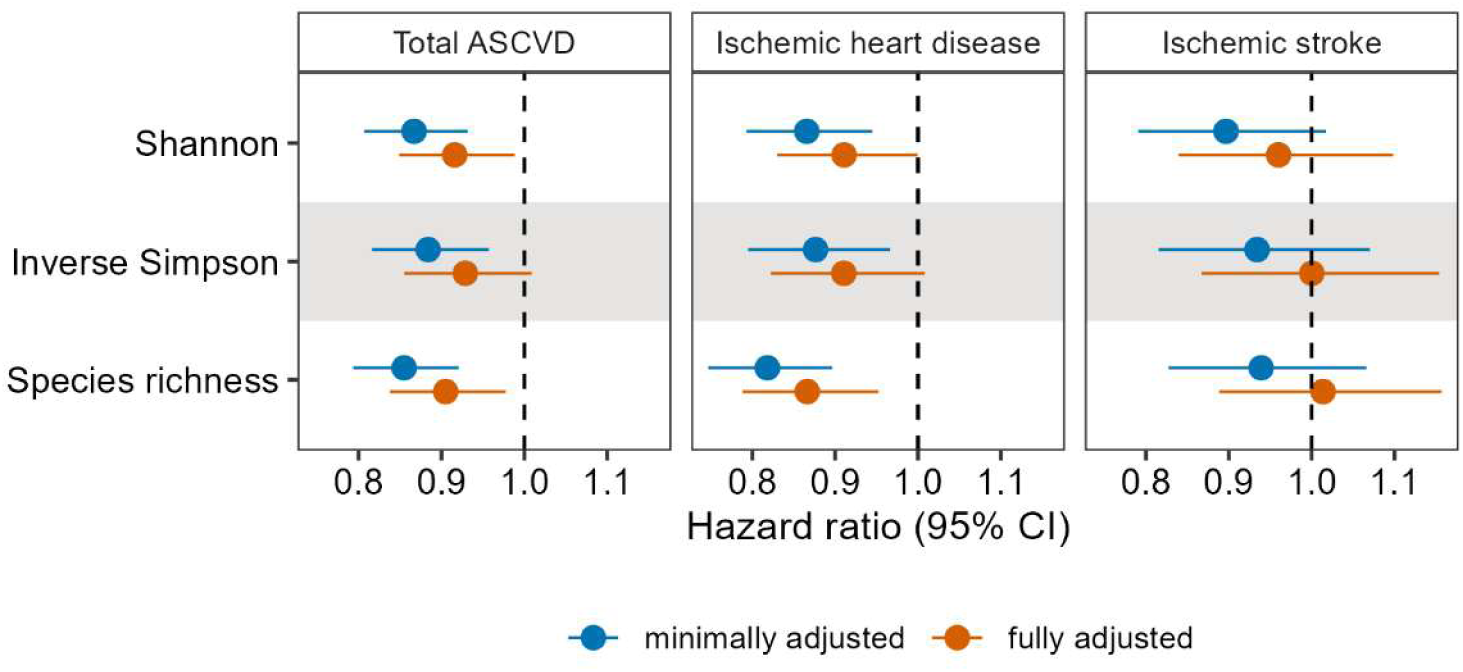
Associations of alpha diversity with incident total ASCVD, IHD, and ischemic stroke events. The forest plot shows hazard ratios and 95% confidence intervals (CIs) from meta-analyses of estimates from SCAPIS, SIMPLER, and MOS for total ASCVD, and from SCAPIS and SIMPLER for IHD and ischemic stroke. Hazard ratios were estimated per 1 standard deviation of Shannon or Inverse Simpson index, and per 100 species increase for richness, with blue color depicting results from the minimally adjusted model, with age as the time scale and adjusted for sex, birth country, and technical variation, and the fully adjusted model (in orange), further including education, physical activity, energy-adjusted carbohydrate, saturated fatty acids, polyunsaturated fatty acids, total energy intake, alcohol intake, cardiovascular medication use, proton-pump inhibitor use, and prevalent diabetes. ASCVD: atherosclerotic cardiovascular disease; IHD: ischemic heart disease; MOS: Malmö Offspring Study; SCAPIS: Swedish CArdioPulmonary BioImage Study; SIMPLER: Swedish Infrastructure for Medical Population-Based Life-Course and Environmental Research.

### Associations of common species with ASCVD

We found 74 and 68 common species associated with risk of ASCVD and IHD, respectively, in the minimally adjusted model (Supplementary Tables 6 and 7, respectively), while none of the species were associated with ischemic stroke after multiple testing correction (Supplementary Table 8). After adjustment for lifestyle, diet, medication use and prevalent diabetes, only eight common species remained significantly associated with IHD (*q*<0.05, Fig. 3, Supplementary Table 7). Direction of associations was consistent across cohorts, with low to moderate heterogeneity (0-65%) in the meta-analyzed estimates. Higher abundances of *S. mutans*, *R. gnavus,* and *Anaerotruncus colihominis* (order *Oscillospirales*) were associated with increased hazard of incident IHD events (Figure 3, Supplementary Table 7). Each doubling of SFEL01 sp004557245 abundance was associated with a 3% lower hazard of IHD events (HR_full_=0.97 [0.95-0.98]). Other species associated with lower IHD hazard included *Blautia_A wexlerae_B* and three uncultured species from the order *Oscillospirales* (*Angelakisella* sp900547385), *Christensenellales* (*Fimadaptatus* sp900553645), and *Lachnospirales* (*Choladousia* sp902363135). Heterogeneity in the estimates was greatly reduced in the fully adjusted model versus the minimal model, except for *Blautia_A wexlerae_B*.

**Figure 3.**
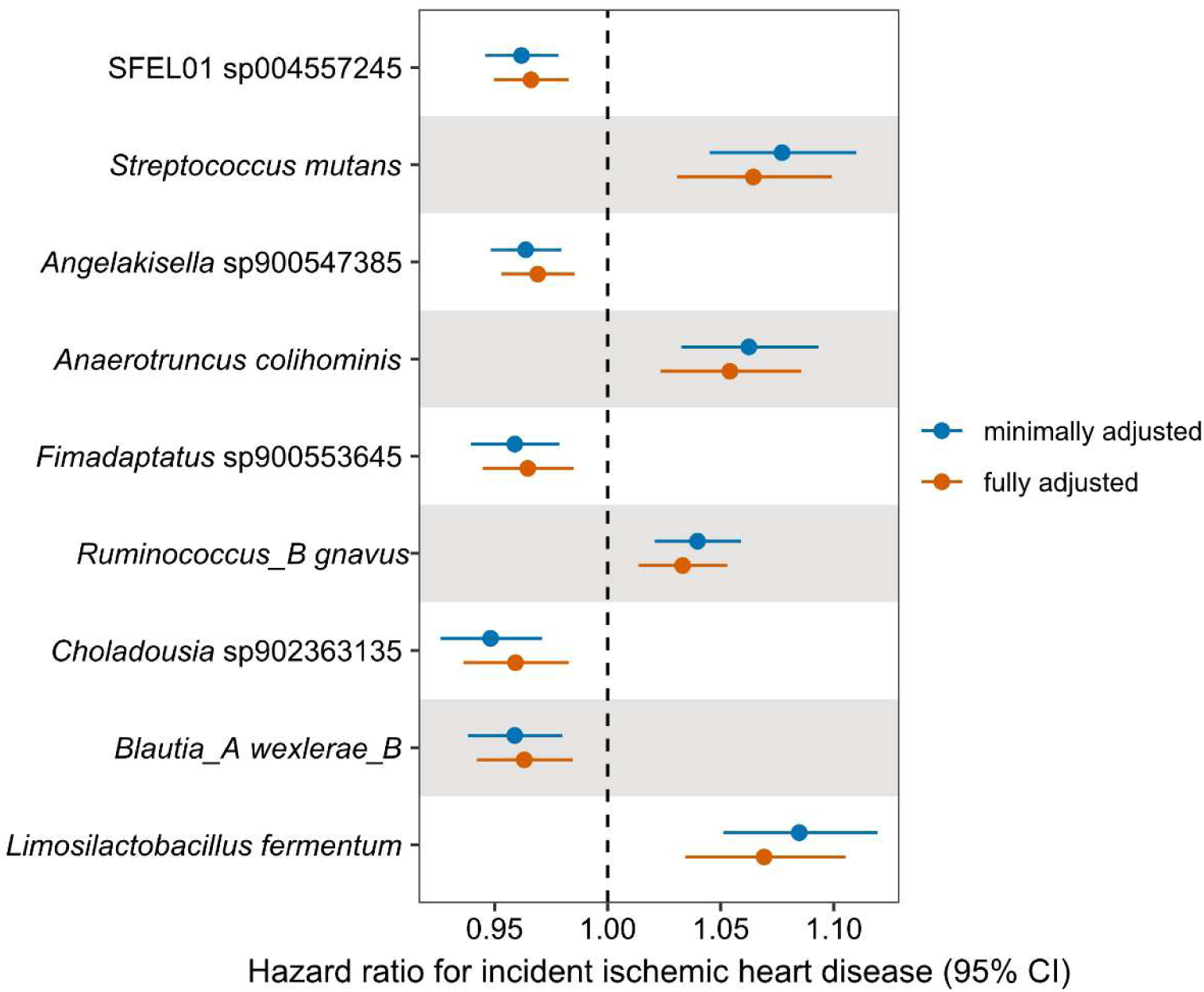
Associations of gut species with incident ischemic heart disease from meta-analysis of two longitudinal Swedish cohorts. The forest plot shows hazard ratios and 95% confidence intervals (CIs) for eight common (>30% prevalence) and one rare candidate species previously associated with coronary artery calcium score in SCAPIS. Hazard ratios were estimated per unit of increase in log2-transformed species relative abundance. The minimally adjusted model (in blue) adjusted for age, sex, birth country, and technical variation, and the fully adjusted model (in orange), further included education, physical activity, energy-adjusted intakes of carbohydrate, saturated fatty acids, polyunsaturated fatty acids, total energy intake, alcohol intake, cardiovascular medication, proton-pump inhibitor, and prevalent diabetes. MOS: Malmö Offspring Study; SCAPIS: Swedish CArdioPulmonary BioImage Study; SIMPLER: Swedish Infrastructure for Medical Population-Based Life-Course and Environmental Research.

### Associations of rare candidate species with ASCVD

We additionally examined candidate species previously associated with CACS in SCAPIS, based on reanalysis of metagenomics data processed using the same pipeline as in the present study (Supplementary Table 9). A total of 155 candidate species with prevalence ranging from 0.01% to 92.1% were associated with CACS (*q*<0.05). For associations with incident events, we meta-analyzed cohort estimates from 80 rare candidate species with prevalence >1% to <30% in SIMPLER and SCAPIS, and >10% to <30% in MOS in relation to total ASCVD and IHD, and from 75 species in relation to ischemic stroke.

Of the 80 rare candidate species, 18 and 19 species, including *Limosilactobacillus fermentum*, *L. oris*, *Ligilactobacillus salivarius*, and *Thomasclavelia ramosa*, were associated with a higher incidence of both total ASCVD and IHD in the minimal model (*q*<0.05) (Supplementary Tables 10 and 11, respectively). However, only one species, *L. fermentum*, remained associated with IHD in the fully adjusted model after multiple testing correction (HR=1.07 [1.03-1.11], *q*=6.1×10^-3^) (Supplementary Table 11). Significant heterogeneity was observed in the meta-analyzed estimates in the fully adjusted model (I^2^ = 83%), with stronger associations observed in SCAPIS than in SIMPLER. No associations were observed in relation to ischemic stroke (Supplementary Table 12).

### Post-hoc associations of S. mutans and L. fermentum accounting for dental health

Two species identified in the primary analyses have been previously associated with dental caries.^32,33^ A higher proportion of individuals with severe periodontitis or ≥5 missing teeth was observed among individuals experiencing IHD events compared to those who remained IHD free, but the proportion for any caries was similar (Supplementary Table 3). In a sensitivity analysis, we adjusted the estimates from the fully adjusted model with either severe periodontitis status, caries status, or the number of missing teeth, and observed similar estimates to the primary analysis (i.e., fully adjusted model) in SCAPIS (Fig. 4).

**Figure 4.**
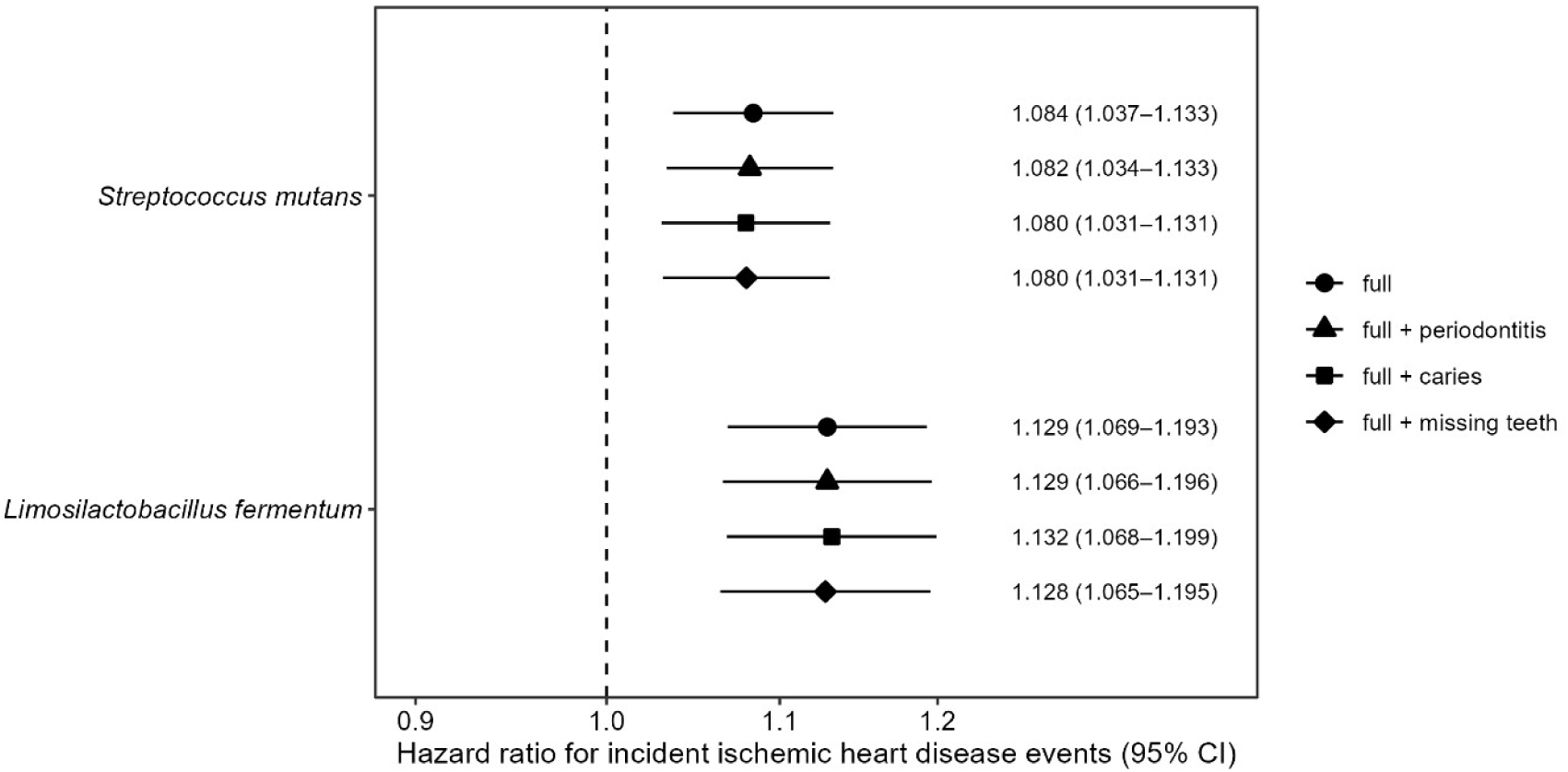
Associations of *Streptococcus mutans* and *Limosilactobacillus fermentum* with incident ischemic heart disease additionally adjusting for dental health measures in the Swedish CArdioPulmonary BioImage Study (SCAPIS; *n* analyzed=9,206, *n* events=216). The forest plot shows hazard ratios and 95% confidence intervals, estimated per unit increase in log2-transformed species relative abundance. The dental health measures included severe periodontitis status (present vs. absent), caries status (any vs. no caries), and the number of missing teeth.

### Sensitivity analysis of microbial species associations

Similar estimates for IHD associations were observed for all species in the BMI-adjusted model, and when excluding individuals with recent antibiotic treatment or those with inflammatory bowel disease (Supplementary Fig. 4, Supplementary Table 13). Including age-at-entry as a stratifying variable in SIMPLER analysis yielded similar associations (Supplementary Fig. 5). Excluding the first two years of follow-up yielded similar estimates, except for *S. mutans*, *A. colihominis*, and *L. fermentum*, which were somewhat attenuated (Supplementary Fig. 4). Further analyses restricting to the first three years of follow-up showed stronger associations for these same species.

Supplementary Fig. 6 showed that results for IHD associations from the primary analyses were strongly correlated to results from other types of transformations applied to the relative abundance data (Spearman’s *r*≥0.96), with slightly lower correlation with results from rank-inverse-normal transformation (for common species, *r_s_*=0.87) or binary presence/absence analysis (for rare candidate species, *r_s_*=0.77).

Species richness as well as other alpha diversity measures were positively correlated with protective IHD species, and negatively with adverse species (Supplementary Table 14). Therefore, to examine whether richness is driving the species-IHD associations, we further adjusted the species associations for richness. Results showed that the estimate was virtually unchanged for SFEL01 sp004557245 after adjustment, while attenuated associations were observed for other protective species such as *Fimadaptatus* sp900553645, as well as the adversely associated *A. colihominis* (Supplementary Fig. 4). Abundances of *S. mutans* and *L. fermentum* were only weakly correlated (absolute *r_s_*≤0.11) with the abundance of protective species or other adverse IHD species or any measures of alpha diversity (Supplementary Table 14).

### Replication in HUNT and FINRISK cohorts

Prevalence of the IHD-associated species was relatively similar in discovery cohorts compared to the replication cohorts, except for *S. mutans,* which was very low (2%) in FINRISK but was 33-43% in other cohorts (Supplementary Table 15), which might indicate a limitation of the measurement in FINRISK, which used shallow-sequencing. A simulation of the effect of low sequencing depth used in FINRISK did, however, not fully explain the low prevalence (Supplementary Methods). Supplementary Table 16 showed results from replication using HUNT (161 events) and FINRISK cohorts (403 events). Analysis in HUNT showed that higher abundance of *S. mutans* was also associated with higher incidence of IHD events (HR 1.06 [1.02-1.11], *P*=0.010). Four of eight species were associated with IHD in the same direction as observed in the discovery cohorts; however, none were statistically significantly associated with incident IHD in the meta-analyzed estimates from the replication cohorts (*P*>0.05).

### Cardiovascular risk factors correlated with richness and IHD-associated species

To investigate the potential causal pathways linked to richness and the IHD-associated species, we examined whether richness and IHD-associated species are correlated with established cardiovascular risk factors in SCAPIS and SIMPLER (Fig. 5). Species richness and most IHD-associated species correlated most strongly with adiposity, hsCRP, GGT, and most lipids, in particular with triglycerides and HDL-C, but not with LDL-C, with comparatively stronger correlations for richness than for individual species. The directions of effect were as expected, given the respective associations with IHD. The strongest correlations (absolute Spearman’s *r*) were 0.22 for richness with triglycerides and adiposity measures in SCAPIS (Fig. 5a; Supplementary Table 17). Of note, *Blautia_A wexlerae_B* and *Choladousia* sp902363135, both from family *Lachnospiraceae*, were only weakly correlated with triglycerides, HDL-C, GGT and hsCRP (absolute *r_s_*<0.10) (Fig. 5, Supplementary Table 17). Correlations with similar direction albeit of lower magnitude were observed for most risk factors in SIMPLER (absolute *r_s_* ≤0.14) (Fig. 5b).

**Figure 5.**
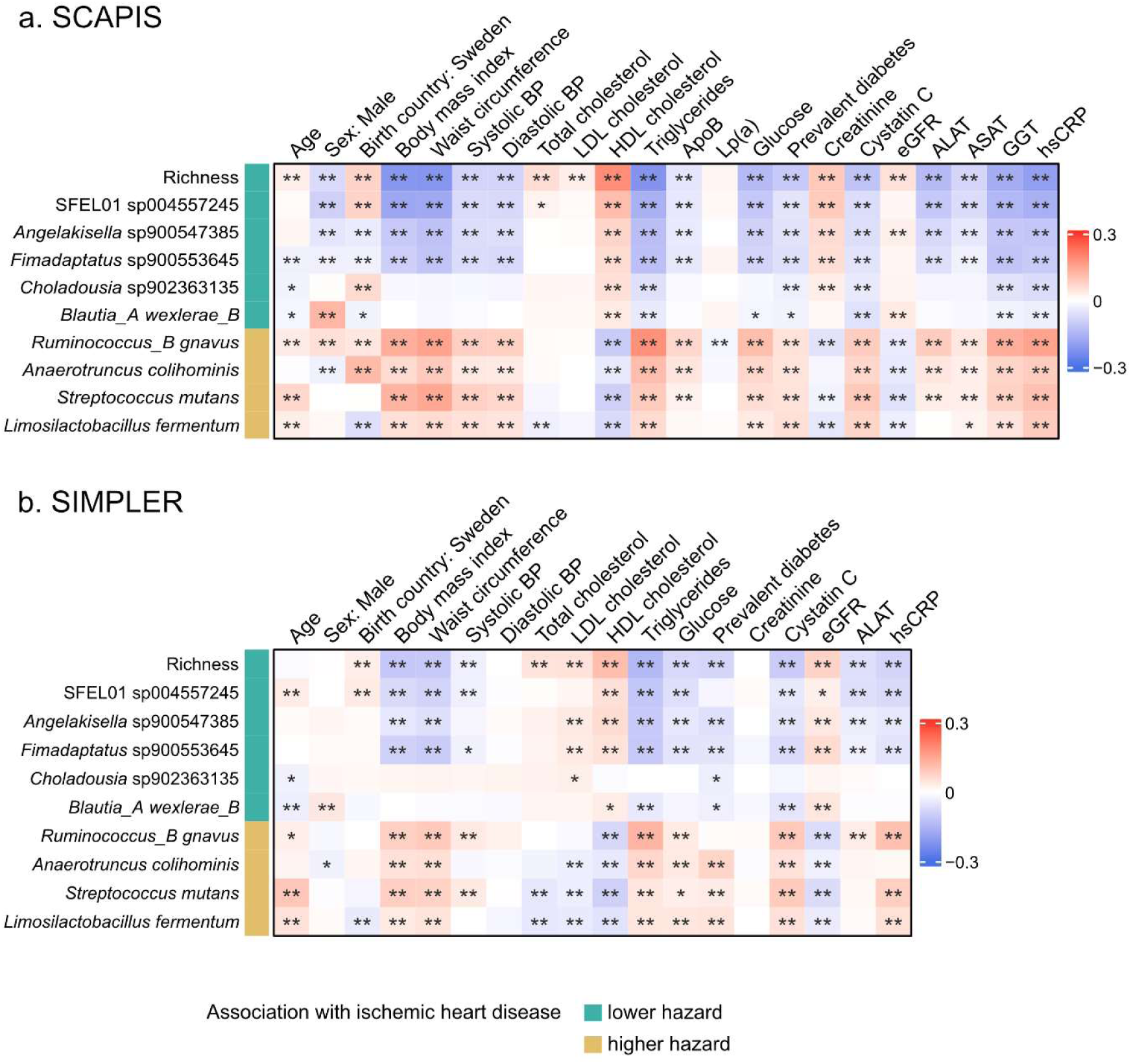
Heatmap of correlations of species richness and nine IHD-associated microbial species with demographic and cardiovascular risk factors in a) SCAPIS (*n* max=8,543) and b) SIMPLER (*n* max=5,485). Spearman’s partial correlations were calculated, with the correlation coefficients shown in red for positive, and blue for negative, and higher intensity indicating a stronger correlation. For age, sex, and birth country, correlations were adjusted for technical variation (plate). For all other risk factors, Spearman’s partial correlations were adjusted for age, sex, and technical variation. The underlying data are available in Supplementary Table 17. Direction of association with the outcome from the main analysis is indicated for each microbiome feature; green shows that the species is associated with a lower hazard of IHD events, and yellow a higher hazard. Asterisks indicate false discovery rate adjusted p-values (*q* values): ** *q*<0.05, * *P*<0.05 but *q*>0.05, and the absence of asterisks indicates *P≥0.05.* ALAT: alanine aminotransferase; ApoB: Apolipoprotein B, ASAT: aspartate aminotransferase; BP: blood pressure; hsCRP: high-sensitivity C-reactive protein; eGFR: estimated glomerular filtration rate; GGT: gamma-glutamyl transferase; HDL: high-density lipoprotein; IHD: ischemic heart disease; Lp(a): Lipoprotein(a); LDL: low-density lipoprotein; SCAPIS: Swedish CArdioPulmonary BioImage Study, SIMPLER: Swedish Infrastructure for Medical Population-Based Life-Course and Environmental Research.

### Metabolites correlated with richness and IHD-associated species

Gut microbial metabolites have been hypothesized to link gut microbiome and ASCVD;^2^ therefore we further examined the correlations of species richness and the IHD-associated species with mass-spectrometry based plasma metabolites in the present study (Supplementary Table 18), with 83 strongest associations ordered by their strength of correlation with species richness summarized in Supplementary Fig. 7. The strongest positive correlation was observed for species richness with 5alpha-androstan-3beta,17alpha-diol disulfate, an androgenic steroid (*r_s_*=0.54), which also correlated with the species in a varying magnitude (absolute *r_s_*=0.08 to 0.45) (Supplementary Table 18). Many metabolites were similarly correlated with richness and IHD-associated species, such as 3-phenylpropionate (hydrocinnamate) and cinnamoylglycine, as well as a few uncharacterized metabolites, were among those with the strongest positive correlations with richness (Supplementary Fig. 7, Supplementary Table 18). Several metabolites previously associated with prevalent cardiometabolic disease were correlated with richness and IHD species. Imidazole propionate correlated negatively with richness (*r_s_*=-0.24) and protective IHD species (r=-0.06 to-0.18), and positively with adverse IHD species (*r_s_*=0.09-0.22). Low to moderate correlations were observed with multiple secondary bile acids (absolute *r_s_≤*0.38); the strongest positive correlation was for isoursodeoxycholate with *R. gnavus* (*r_s_*=0.38), and the same bile acid was negatively correlated with species richness (*r_s_*=-0.37). In comparison, abundances of *S. mutans* and *L. fermentum* were not correlated or only weakly correlated with secondary bile acid metabolites (*r_s_*<0.1). Additionally, *S. mutans* abundance correlated negatively with vitamin A metabolites and ergothioneine, and positively with the omeprazole metabolite and several tobacco metabolites (Supplementary Table 18, Supplementary Fig. 7).

## DISCUSSION

We present a large prospective study assessing the association between the gut microbiome and incident ASCVD, including 17,216 community-dwelling participants with 769 ASCVD events over a median follow-up of 6-8 years. We identified four bacterial species that were adversely associated with IHD after extensive adjustment for confounders, including the oral bacteria *S. mutans* and *L. fermentum*. Furthermore, gut microbial diversity and five species were inversely associated with risk of IHD.

We found that the gut abundance of the oral species *S. mutans* and *L. fermentum*, known to be involved in dental caries when present in the mouth, was positively associated with IHD in the Swedish cohorts, with similar effect estimates in the HUNT study. These findings corroborate our earlier cross-sectional analysis of the microbiome composition in relation to image-based measures of coronary atherosclerosis in SCAPIS.^12^ Gut abundances of *Streptococcus* and other oral taxa have been repeatedly reported to be associated with prevalent cardiovascular disease,^34–39^ and in a recent prospective study with new-onset hypertension.^14^ Oral disease has been associated with increased ASCVD risk,^28,33^ with proposed mechanisms including increased translocation of oral bacteria and/or bacterial components from the oral cavity to the gut and/or to circulation due to poor oral health, as well as modulation of local and systemic inflammation by the oral bacteria.^33,40^ It has also been suggested that an increased abundance of oral species in the gut is a marker for microbial imbalance (“dysbiosis”) and depletion of commensal bacteria.^41^ Our analysis strategy accounted for medications related to such depletion, and models adjusted for diversity showed virtually unchanged estimates, making this explanation less likely. We also explicitly accounted for several measures of oral health and observed similar estimates, indicating that adverse associations for these species cannot be fully explained by poor oral health. A recent study showed that the same strain of *S. mutans* was found in the oral cavity and the gut, but exhibited different gene expression and predicted functions, suggesting that *S. mutans* was able to adapt to the gut environment.^42^ We observed somewhat attenuated associations for *S. mutans* and *L. fermentum* in the analysis excluding the first two years of follow-up, which could indicate reverse causation, temporal instability of the exposure, or a chance finding. However, we consider reverse causation unlikely as we have excluded individuals with prior ASCVD events in our analysis and adjusted for cardiovascular medications. Temporal instability might explain the diminished effects of an exposure as the follow-up period lengthens.^43^ Members of *Lactobacillales*, such as the genus *Streptococcus*, have been previously reported to largely fluctuate within individuals over time.^44^

Enrichment of *R. gnavus,* a prevalent Gram-positive bacterium (assigned *Mediterraneibacter gnavus* in GTDB R232) was associated with increased IHD risk in the present study, consistent with several previous studies. Nooij and colleagues showed that *R. gnavus* is particularly enriched in ASCVD patients from 86 studies.^45^ In the HELIUS cohort (9.5 y follow-up, 129 events) *R. gnavus* group spp. was associated with an increased risk of major adverse cardiovascular events in the age-adjusted model, although it was no longer associated in multivariable analysis.^45^ *R. gnavus* is further reported to correlate with many traditional CVD risk factors, such as excess adiposity^46,47^ and prevalent,^45^ as well as incident type 2 diabetes.^48,49^ *R. gnavus* might affect IHD risk by modulating the levels of microbial-derived metabolites such as secondary bile acids^50^ and imidazole propionate,^51,52^ which were also observed in the present study. Another species associated with IHD risk, *A. colihominis*, showed a similar pattern of associated metabolites to *R gnavus* and has been shown to be enriched in patients with acute coronary syndrome.^39^ While *A. colihominis* and *R. gnavus* have been reported to produce SCFAs, in SCAPIS data, their abundances were correlated negatively with dietary fiber intakes and positively with inflammatory marker CRP. *A. colihominis* and some strains of *R. gnavus* have been reported to utilize mucin as a nutrient source.^53,54^ A habitually low fiber diet was shown to promote expansion of mucin-degraders, and lead to increased inflammation,^55^ which potentially explains the adverse association for these species in the present study. Abundance of both species was also enriched in those with recent antibiotic use.^56^ We also observed that further adjustment for microbial diversity or exclusion of individuals with recent antibiotic use attenuated the associations of both species. This suggests that the increased abundance of *R. gnavus* and *A. colihominis* might also simply reflect a microbial community with a reduced diversity. Gut enrichment of inflammatory-associated taxa, such as *R. gnavus*, has been suggested to characterize a microbial community with low diversity,^57^ although the directionality of causality is unknown.

We observed an inverse association between gut microbial diversity and risk of IHD. Previous evidence for an association of microbial diversity and ASCVD is conflicting; most case-control studies^6,37,38,58^ reported no associations, while a limited number of cross-sectional analyses in individuals without prior CVD showed that microbial diversity was inversely associated with CVD risk score^59,60^ and with subclinical atherosclerosis in SCAPIS.^12^ The few available prospective analyses also reported no associations, but they were likely insufficiently powered (≤254 events).^13–15^ High microbial diversity is generally regarded as beneficial for health, given the low microbial diversity observed in diseased states, such as inflammatory bowel disease.^61^ Westernization, often characterized by low fiber and high saturated fat content in the diet, has been associated with lower microbial diversity, with concomitantly reduced microbial capacity to degrade complex carbohydrates, resulting in lower production of short-chain fatty acids,^62–64^ involved in host physiological processes also related to the pathogenesis of ASCVD, such as lipid metabolism and immune system modulation.^2,65^ Antibiotic use, which severely reduces microbial diversity, has also been linked to increased CVD events in a few studies.^66,67^ In the present study, individuals with higher microbial diversity at baseline tend to have healthier lifestyles and better cardiometabolic health compared to those with less diverse gut microbes, consistent with previous cross-sectional studies.^57,59,68,69^ The association of gut microbial diversity with the risk of IHD in the present study remained after extensive adjustment of demographic and lifestyle factors, medication use, and prevalent diabetes, known to affect both gut microbiome and ASCVD risk, which suggests that microbial diversity may also affect ASCVD risk through another pathway that is independent of these factors. A highly diverse ecosystem is thought to be stable and resilient; a diverse microbial community is more resistant to perturbations and would recover faster from perturbation than a community with lower diversity, due to high functional redundancy.^70–72^ Impaired resilience against perturbations therefore may have detrimental consequences for host health. Additional research is needed to elucidate the pathways through which microbial diversity confers protection against ASCVD.

In this study, several taxa are associated with a lower risk of IHD. SFEL01 sp004557245 and *Fimadaptatus* sp900553645 belong to the order *Christensenellales*, whose members have been linked to cardiometabolic health and butyrate-producing capacity. In a recent study, SFEL01 sp004557245 was identified as having *buk*, one of the terminal genes for butyrate production from complex polysaccharides, encoding butyrate kinase.^73^ Depleted abundance of members of *Christensenellales* has previously been reported to be associated with higher cardiovascular risk (estimated by CVD risk scores) in individuals without CVD.^13,60,74^ We further investigated their associations with plasma metabolites, revealing many shared, correlated metabolite associations with species richness. We also recently found the same genetic variant to be associated with both species richness and SFEL01 sp004557245,^17^ leading us to suspect that the abundance of this species might be a marker of microbial diversity. However, adjusting for species richness and excluding antibiotic users showed virtually unchanged estimates for SFEL01 sp004557245, suggesting that its abundance is not merely a predictor of high diversity. Associations for other protective species, *Blautia_A wexlerae_B*, and uncultured species *Fimadaptatus* sp900553645, *Choladousia* sp902363135 and *Angelakisella* sp900547385, were slightly attenuated after adjusting for species richness or antibiotic use. These uncultured species have not been previously described in relation to ASCVD or its risk factors, except for *Angelakisella* sp900547385 whose abundance was inversely associated with visceral fat^75^ and 24-hour blood pressure.^76^ Although still limited, there is growing evidence linking *Blautia wexlerae* (NCBI taxonomy for *Blautia_A wexlerae_B*) to obesity and diabetes.^77–79^ *B. wexlerae* abundance was inversely associated with prevalent obesity and type 2 diabetes in Japanese individuals,^78^ and in two different experiments, interventions with *B. wexlerae* seemed to protect against obesity and diabetes in murine model.^78,79^ Although *B. wexlerae* lacks the ability to produce propionate and butyrate, it can produce succinate, lactate, and acetate,^78^ which can be used by other bacteria to produce propionate or butyrate. Such cross-feeding interactions may also be important for cardiovascular health and warrant further investigation.

Strengths of our present analyses include a prospective design, a large sample size, register-based ascertainment of outcomes, harmonized high-resolution microbiome characterization, and extensive adjustment for confounders, with prioritized species that are robust to variation in data transformation methods. Firstly, the prospective design substantially reduced the risk of reverse causation compared with previous studies that largely relied on case-control designs. Secondly, the number of ASCVD events included in this study is by far the largest owing to linkage to national patient and death registers, enabling us to obtain detailed information about diagnoses, hospitalizations, and causes of death, and to reduce the risk of outcome misclassification^80^. Third, shotgun metagenomic sequencing and a harmonized bioinformatic pipeline across cohorts enabled species-level resolution. Previous studies have used low-resolution 16S rRNA data^6,13,14^ or shallow metagenomic sequencing^15^ and thus were only able to report findings at genus level or higher. Fourth, the extensive inclusion of confounding factors is important because environmental factors such as diet and medications shape the composition of an individual’s gut microbiome and are also linked to cardiovascular risk.^8–11^ However, previous studies have not fully accounted for these factors, as well as other lifestyle factors such as smoking, physical activity, and presence of diabetes.

When we adjusted only for demographic and technical variation, as in previous studies, 68 common and 19 rare species were associated with IHD; after further controlling for differences in lifestyle, medication use, and prevalent diabetes, only eight common and one rare species remained. This underscores the importance of rigorous control of confounding in causal investigations of the microbiome and disease risk, and helps prioritize species for further functional characterization and as potential targets for intervention. Additionally, the harmonization of confounders appears important when including data from multiple cohorts. Heterogeneity in associations among most species was substantially reduced in the fully adjusted model, suggesting that after accounting for differences attributable to lifestyle and medications, the associations are more consistent across cohorts. To note, our model specification assumes that associations of microbial species with ASCVD have a similar confounding structure, which may not hold across all tested microbial species; however, this modeling approach also allows us to test a large number of species and provides greater flexibility in testing our assumptions through multiple sensitivity analyses. Some studies controlled for differences in BMI statistically or by matching, instead of adjustment for differences in diet or physical activity.^38,60,81^ However, BMI might also lie in the causal pathway between the gut microbiome and ASCVD,^82,83^ and importantly, adjusting for BMI but not diet or physical activity assumes that the influence of these factors on the gut microbiome and ASCVD risk is solely through BMI modification. Nevertheless, our results showed that further adjustments for BMI showed virtually unchanged estimates, likely because diet and physical activity, already adjusted for in the analysis, are strongly correlated with BMI.

Several limitations also need to be considered. Owing to the observational nature of the study, we cannot offer definitive conclusions regarding the causal relationship between the gut microbiome and ASCVD. Despite our efforts to control for confounding and the availability of extensive linkage with registers, residual confounding due to measurement errors cannot be ruled out. We tried to limit this as much as possible, for example, by capturing proton-pump inhibitors use based on their detected metabolites in plasma, as these drugs can be both prescribed and available over the counter. We also attempted to replicate our findings in external cohorts; however, meta-analyzed estimates from HUNT and FINRISK cohorts showed largely non-replicated associations, except for *S. mutans*, which was adversely associated with IHD in HUNT participants and was not reliably measured in the shallow-sequenced FINRISK. While these results suggest that these species may not be truly associated with ASCVD, we cannot rule out methodological differences and the much longer follow-up duration in FINRISK contributing to the non-replication. The low prevalence of some species might have further limited the statistical power to detect associations. Inconsistency in published findings remains a concern in microbiome research,^84^ partly due to a lack of standardized protocols for sample collection, DNA extraction and sequencing, and bioinformatics pipeline, which can substantially influence the observed microbial composition.^85–87^ Despite these limitations, our study could offer insights on the importance of ensuring sufficient power (i.e., having many events) by including more cohorts, ideally with gut microbiome data processed using a standardized bioinformatic pipeline, and controlling for important covariates such as diet, lifestyle and medication use, in microbiome-disease studies aiming for causal inference. In this study, the fecal metagenomic data was only available at baseline, assuming that all microbial species remained stable throughout the follow-up period. Investigations using repeated measurements of the gut microbiome and atherosclerosis progression would provide stronger evidence of causality between the gut microbiome and ASCVD.

Collectively, our data support the role of the gut microbiome in the pathophysiology of ASCVD. Our present analysis provides evidence for the protective effect of higher microbial diversity on the long-term risk of ASCVD and shows that low microbial diversity can be present years before the disease occurs, supporting the temporality of the association. Our data also add to the evidence that gut enrichment of oral taxa *S. mutans* and *L. fermentum*, and inflammatory-associated *R. gnavus,* is likely harmful, while enrichment of two *Christensenellales* species seems to be protective. Furthermore, our study identifies several microbial features as targets to prioritize in future studies to confirm their causal roles.

## Data Availability

Summary data supporting all figures and analyses are available as Supplementary Tables accompanying the manuscript. The pseudonymized SCAPIS, SIMPLER, and MOS metagenomics and phenotype data supporting the conclusions of this article were provided by the respective central data offices and are not shared publicly due to confidentiality and ethical restrictions. Data will be shared only after permission from the Swedish Ethical Review Authority (https://etikprovningsmyndigheten.se) and from the data access board (https://www.scapis.org/data-access, https://www.simpler4health.se, and https://www.malmo-kohorter.lu.se/malmo-offspring-study-mos). Similarly, access to pseudonymized data from the replication cohorts the HUNT study and FINRISK requires approval from the respective ethical and data access boards. Publicly available software was used for the analyses described in the Methods section. The source code will be made available on Github at https://github.com/MolEpicUU/microbiome_ascvd.

## Author contributions

K.P., T.F., K.F.D., U.H., S.C.L planned and designed the study, including replication in external cohorts; T.F., M.O.-M, K.M., J.Ä., G.E., J.G.S., O.M. collected data for discovery cohorts; K.P. performed the statistical analyses; K.P. and K.F.D. performed analyses with metabolomics data; M.D.-V. and S.S.-B performed reanalysis of gut microbiome and coronary artery calcium score; E.C., O.K. analyzed replication data. C.O., K.H. provided replication data from the HUNT study; T.N., A.S.H., V.S. provided replication data from the FINRISK study; P.P. and D.J. provided dental registry data. K.P. wrote the first draft of the manuscript; T.F., S.C.L, G.B., K.F.D, S.Y., U.H., R.H., M.M., M.O.-M, J.Ä., J.G.S., G.M., C.G.V., K.M., D.G., O.K., V.S., D.J., P.P., M.D.-V., S.S.-B, K.H., and R.K. contributed to the editing of the initial manuscript draft. All authors provided intellectual input, reviewed, and approved the final version of the manuscript.

## Cohort acknowledgment

This research has been conducted using the Swedish CArdioPulmonary bioImage Study (SCAPIS) Resource, under petition number 774 and 423. The main funding body of SCAPIS is the Swedish Heart and Lung Foundation. The study is also funded by the Knut and Alice Wallenberg Foundation, the Swedish Research Council, VINNOVA (Sweden’s Innovation agency), the University of Gothenburg and Sahlgrenska University Hospital, Karolinska Institutet and Region Stockholm, Linköping University and University Hospital, Lund University and Skåne University Hospital, Umeå University and University Hospital, Uppsala University and University Hospital. We would like to acknowledge the help of Biobank Sweden and the local biobank facilities for their services in handling of biological samples and biobanking. This research also used data from the National Research Infrastructure SIMPLER (www.simpler4health.se), supported by the Swedish Research Council (grant number 2017-00644, 2017-06100, and 2021-00160) and Stiftelsen Olle Engkvist Byggmästare. The Trøndelag Health Study (HUNT) is a collaboration between HUNT Research Center (Faculty of Medicine and Health Sciences, Norwegian University of Science and Technology NTNU), Trøndelag County Council, Central Norway Regional Health Authority, and the Norwegian Institute of Public Health. We thank all FINRISK 2002 study participants for their generous participation in Finnish health research.

The computations and data handling were enabled by resources in project simp2023022 and sens2019512 provided by the National Academic Infrastructure for Supercomputing in Sweden (NAISS) at UPPMAX, funded by the Swedish Research Council through grant agreement no. 2022-06725. We would like to acknowledge the Karolinska Institute Biobank for their services regarding DNA extraction. The data used for the replication analysis in FINRISK were obtained from THL Biobank (study number: THLBB2025_17). We thank the Health and Social Data Permit Authority Findata for providing the health register data (permit number: THL/3273/14.02.00/2025) and wish to acknowledge CSC-IT Center for Science, Finland, for computational resources.

